# Precision therapy with ampreloxetine for neurogenic orthostatic hypotension in multiple system atrophy

**DOI:** 10.1101/2025.08.12.25332833

**Authors:** Roy Freeman, Horacio Kaufmann, Italo Biaggioni, Valeria Iodice, Jens Jordan, Ross Vickery, Tadhg Geurin, Matthew J. Kmiecik, Lucy Norcliffe-Kaufmann

## Abstract

**Objective:** A pre-specified subgroup analysis of ampreloxetine (oral, 10 mg/once-per-day) for neurogenic orthostatic hypotension (nOH) in patients with multiple system atrophy (MSA).

**Background:** The degeneration of the central autonomic network underlies neurogenic orthostatic hypotension (nOH) in patients with multiple system atrophy (MSA), which leads to disability. The pathology shows relative sparing of the peripheral autonomic neurons. Despite adherence to treatment many patients remain symptomatic. Ampreloxetine is a novel, long-acting, selective, norepinephrine re-uptake inhibitor that allows for once daily dosing to precisely target residual peripheral autonomic neurons. Based on the hypothesis that MSA patients would be most responsive to ampreloxetine, we performed a pre-specified subgroup analysis of the MSA cohort.

**Methods:** REDWOOD (NCT03829657) was an IRB-approved, phase 3, placebo-controlled, randomized withdrawal trial conducted at 76 international clinics. The trial had a 16-week open-label period with responders continuing into a double-blind 1:1 randomized 6-week withdrawal phase. Outcome measures included symptom burden captured on the 10-item OH-Questionnaire (OHQ), blood pressures, and catecholamine assays. The pre-specified subgroup analysis included all randomly assigned MSA patients.

**Results:** In total, 73 MSA patients entered the program. Forty (61%) met enrichment criteria and went on to randomize. At the end of the open-label phase, their average OHQ symptom assessment (OHSA) composite score improved by 2.6 points from the pre-treatment baseline values. At week 6 of randomization, symptoms remained stable in the ampreloxetine group but worsened on placebo (mean difference OHSA composite score: -1.6 points; p=0.0056). Walking for a short time showed similar changes favoring ampreloxetine (-2.0 points; p=0.0147). Standing 3-minute blood pressure remained unchanged from open-label in the ampreloxetine group (systolic: 5.6±4.14; diastolic: 3.7±2.89 (SE) mmHg), but dropped in the group withdrawn to placebo (systolic: -10.0±4.45, diastolic -6.0±3.09 mmHg). The catecholamine profile showed target engagement of norepinephrine transporter-inhibition by ampreloxetine. There were no safety concerns nor observed increases in supine blood pressure.

**Conclusion:** If the ongoing phase 3 study confirms the safety and efficacy, ampreloxetine would be the first example of a precision medicine approach for the treatment of nOH in MSA.

## INTRODUCTION

Multiple system atrophy (MSA) is a rare, sporadic neurodegenerative synucleinopathy characterized by autonomic failure, parkinsonism and cerebellar features in varying combination. ^1^ Prevalence estimates range from 3.1 to 12.4 per 100,000 in the US population. ^2^ The disease is rapidly progressive and usually leads to death 5-8 years after the onset of symptoms. Few patients survive for more than 10 years. ^3^

Pathologically, MSA is defined by the deposition of insoluble aggregates of misfolded alpha-synuclein in the nervous system. The hallmark of the disease is the deposition of alpha-synuclein in glial cytoplasmic inclusions, in contrast to the Lewy bodies seen in neurons of autopsied cases of patients with Parkinson disease (PD). ^4^ The gross pathology of MSA involves degeneration of the striatonigral system, cerebellum, pons, olives and central autonomic network with relative sparing of the peripheral autonomic neurons. ^1 5^ This manifests in patients having normal venous plasma norepinephrine (NE) levels, compared to other Lewy body diseases (including PD and pure autonomic failure [PAF]). ^5 6^ where there is substantial degeneration of peripheral autonomic neurons. ^7^

Degeneration of the central autonomic network in MSA underlies neurogenic orthostatic hypotension (nOH) in patients with MSA, which leads to physical deconditioning, functional decline, frequent falls, disability, dependency, and reduced quality of life. Furthermore, nOH is associated with an increased risk of cognitive impairment and early death from all-cause mortality. ^8 9 10 11^ Up to 80% of MSA patients will develop neurogenic orthostatic hypotension (nOH) in their lifetime. Despite adherence to non-pharmacological interventions and the available pharmacological treatments, many patients remain symptomatic. A pressing need exists for additional effective pharmacotherapies.

The implementation of precision medicine—the right drug for the right patient administered in the right dose—has advanced clinical care and improved outcomes in several medical disciplines, most notably cancer. ^12^ To date, apart from in rare enzyme deficiencies, ^13 14^ this approach has not been widely adopted in autonomic neurotherapeutics, particularly in the neurodegenerative synucleinopathies where the therapy could be tailored to the pathophysiological lesion. ^15^

Ampreloxetine is a novel oral, selective, long-acting, norepinephrine reuptake inhibitor that reaches a steady state of NE transporter inhibition and allows for once daily dosing to precisely target residual peripheral autonomic neurons. ^16^ Phase II studies showed 10 mg/day oral ampreloxetine raised venous plasma norepinephrine levels in patients with nOH caused by all synucleinopathies. ^17^ The mechanism of action, however, is best suited to MSA patients with a central pattern of degeneration.

Two back-to-back phase 3 trials of ampreloxetine for the treatment of nOH in patients with synucleinopathies (SEQUOIA, NCT03750552 and REDWOOD, NCT03829657) were conducted. In these trials, a prespecified hypothesis was that MSA patients with relative sparing of peripheral autonomic neurons, would be most responsive to ampreloxetine. Randomization was stratified by disease type in both trials and a pre-specified MSA sub-group analysis was performed to determine the effect of ampreloxetine on symptoms, activities, and cardiovascular autonomic measures. Here we report the results of that analysis.

## METHODS

### STUDY DESIGN AND OVERSIGHT

SEQUOIA and REDWOOD were sequential multi-center, double-blind, placebo-controlled, randomized trials of a 10 mg/day single oral dose of ampreloxetine conducted at 76 clinical sites in 19 countries (**eTable 1**). Eligible patients were screened and enrolled in the 4-week run-in double-blind parallel-group, randomized, controlled trial (SEQUOIA) and rolled over into REDWOOD, which included a 16-week open-label steady-state enrichment period, with responders continuing into a double-blind, randomized withdrawal period (RW; 6 weeks).

The first patient in SEQUOIA began receiving ampreloxetine or placebo in January 2019 and the first patient entered REDWOOD in February 2019. The last patient began receiving ampreloxetine or placebo in June 2021 and the final patient entered REDWOOD in September 2021. Potential candidates were identified at trial sites by trained physicians selected for their expertise in autonomic and/or movement disorders. The trials were designed by the sponsor (Theravance Biopharma) and an advisory board of independent key opinion leaders. Theravance Biopharma provided ampreloxetine and matching placebo tablets, monitored the trial sites, performed the laboratory tests, and analyzed the data.

### PARTICIPANTS

The trials enrolled participants ≥30 years of age with PD, PAF, or MSA and symptomatic nOH. The pre-specified subgroup analysis was performed in the patient population that entered the trial with an established diagnosis of MSA according to consensus criteria at the time of the trial (**Figure 1**). ^18^ In addition, the entry criteria included: 1) diagnosis of nOH defined as a sustained fall in blood pressure of ≥20 or ≥10 mmHg (systolic/diastolic) within 3 minutes of upright tilt to ≥60°; ^19^ 2) a Montreal Cognitive Assessment (MoCA) score >23 points, ^20^ and 3) plasma NE levels ≥100 pg/mL after 30 minutes seated. Other inclusion criteria included a symptom severity score ≥4 points in dizziness/lightheadedness on the validated Orthostatic Hypotension Questionnaire (OHQ, item #1, range 0 – 10, with a 0 representing no symptoms and a 10 worst possible symptoms). ^21^ Patients with severe diabetic neuropathy were excluded (for full entry criteria see **eMethods**).

**Figure 1:**
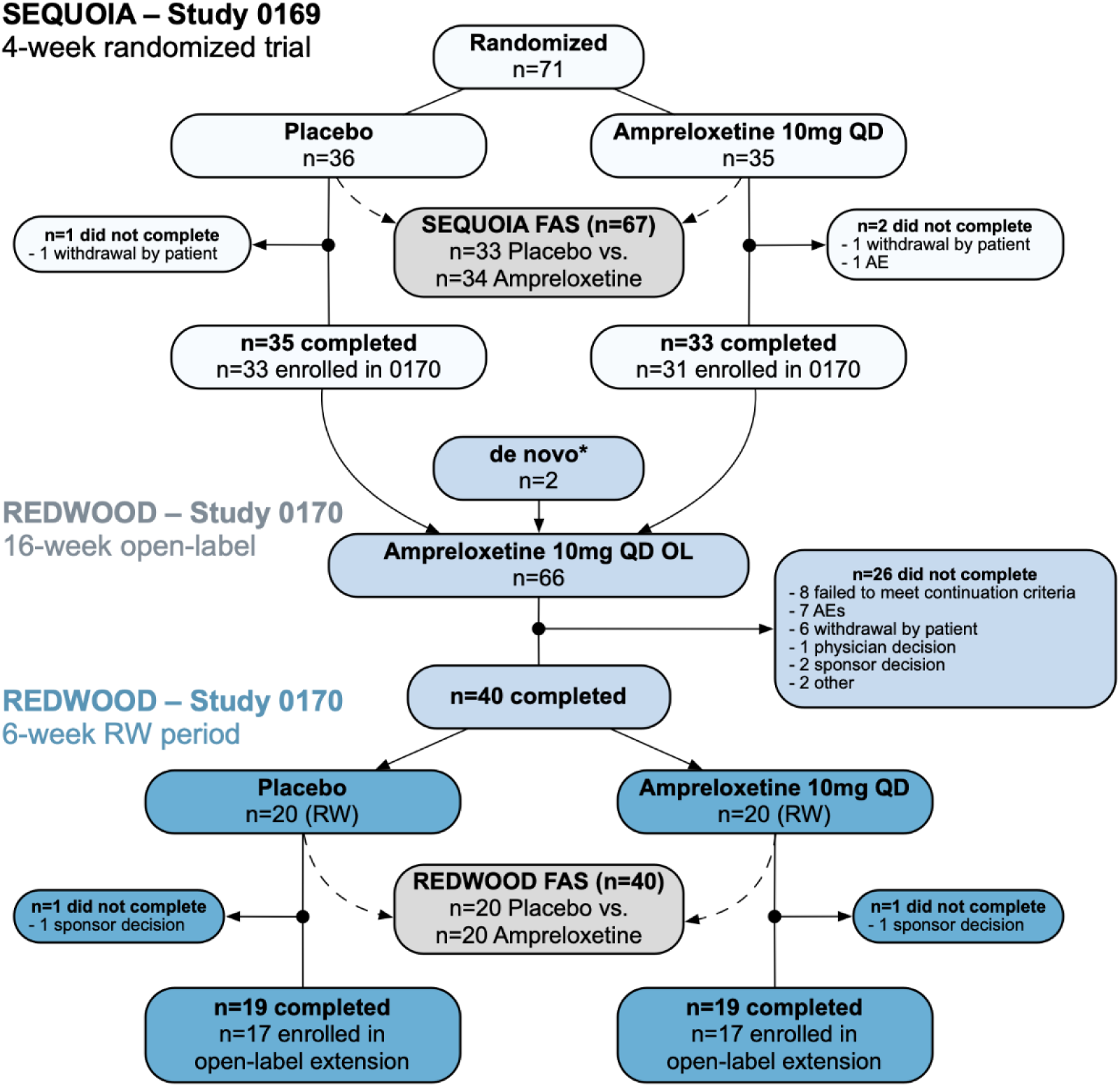
CONSORT Flow Diagram. Patient flow for the MSA subgroup enrolled in the 4-week randomized control trial (SEQUOIA) and 22-week randomized withdrawal study (REDWOOD). After SEQUIOA did not meet the primary endpoint, REDWOOD was terminated early by the sponsor. Pre-specified subgroup analysis of the REDWOOD RW included 40 MSA participants that randomized withdrawal phase, all of which completed. *Both de novo patients were terminated early in the open-label phase by the sponsor due to closure of the trial and therefore were not included in the analysis set. AE = adverse event; FAS = full analysis set; OL = open-label; RW = randomized withdrawal, QD = once daily.

Investigators were required to submit a written clinical narrative to support an MSA diagnosis and stage of the disease at the time of screening. Eligibility was confirmed by adjudication of an Enrollment Steering Committee consisting of 3 independent neurologists with extensive clinical and research experience in autonomic failure and movement disorders. The Committee was required to confirm: 1) The diagnosis of MSA; and 2) the presence of symptomatic nOH. If discordance occurred the Committee Chair had final determination. Patients with probable and possible MSA with the cerebellar and parkinsonian phenotype were included in the trial. After screening, participants were withdrawn from other pressor agents (including: midodrine, droxidopa). Fludrocortisone was allowed at a stable-dosing regimen, initiated at least 7 days prior to enrollment.

### PROCEDURES

**Figure 2** depicts the trial design and visit schedule. Approved participants who met full entry criteria, were first randomized 1:1 to a once daily 10 mg ampreloxetine dose or matching placebo. The randomization sequence was generated using permuted blocks and was stratified by disease type. A central interactive randomization and trial supply management system was utilized to assign participants to treatment arms and manage the dispensing of investigational product, ensuring concealment of the allocation sequence and maintenance of the blind for participants, investigators, and study staff. Neurological scores from the Unified Multiple System Atrophy Rating Scale (UMSARS) were obtained to determine disease stage. ^22^ Participants were instructed to take a single oral 10 mg dose of ampreloxetine or placebo with 8 ounces of water at the same time each morning. No food or fluid restrictions were imposed. Education on non-pharmacological strategies to manage nOH was provided, including maintaining adequate salt/water intake, sleeping with the head of the bed elevated at the same angle, and the use of physical counter maneuvers (i.e., muscle tensing) to avoid impending syncope. Those that successfully completed the 4-week RCT phase in SEQUOIA, with 80% trial medication compliance, were rolled over into REDWOOD, which started with a 16-week open-label steady-state period. The open-label steady-state period of REDWOOD included an enrichment design with participants required to show symptomatic benefit compared to baseline at two enrichment continuation points: 1) at week 4, which required a ≥2 points improvement in dizziness/lightheadedness (OHQ item#1) ^21^ over pre-treatment baseline values; and 2) at week 16, which required participants to have a dizziness/lightheadedness score of ≤7, which allowed room for worsening following withdrawal to placebo. Randomization was stratified by diagnosis using a computer-generated randomization sequence to allow similar numbers of MSA patients across both arms. To minimize dropouts, 1 rescue dose of midodrine per week (10 mg max) was allowed during the 6-week RW segment. Due to the COVID-19 pandemic, the protocol was amended to allow for remote home visits conducted by a trained nurse under the supervision of the treating investigator, who appeared via secure audio-visual telemedicine link.

**Figure 2:**
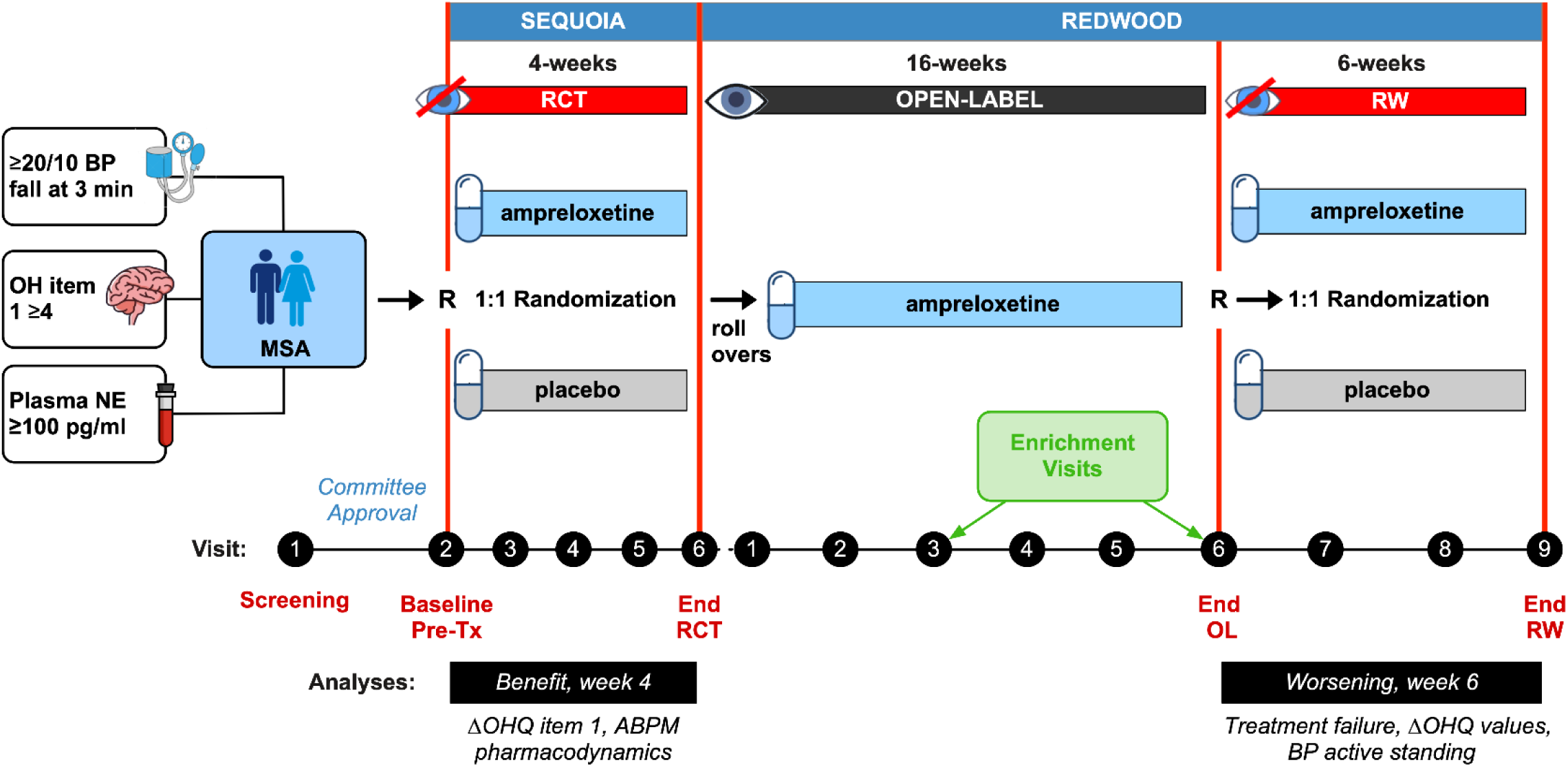
Study Design. The pre-specified subgroup analysis was performed in participants enrolled with a diagnosis of MSA and symptomatic nOH, approved by an Enrollment Steering Committee, who met the cut off for preserved peripheral (sympathetic) autonomic neurons (plasma NE ≥100 pg/mL). Approved participants were enrolled in a 4-week double-blind, placebo-controlled, randomized controlled clinical trial of ampreloxetine (SEQUOIA). After completion, participants rolled over into REDWOOD, which included a 16-week open-label period, with 2 enrichment points (week 4 and week 16). Participants that met open-label enrichment criteria underwent randomized withdraw with 1:1 allocation to placebo or ampreloxetine for 6 weeks. The analysis of the efficacy outcome measures were performed at the end of each double-blind phase. SEQUOIA was designed to show benefit of ampreloxetine over placebo using the OHQ item 1. REDWOOD was designed to show worsening after withdrawal to placebo in a composite treatment failure endpoint (1 point worsening of OHQ item 1 and a 1-point worsening of global patient impression of severity). BP = blood pressure; OHQ item 1 = Orthostatic Hypotension Questionnaire item 1 (dizziness/lightheadedness); NE = venous plasma norepinephrine; MSA = multiple system atrophy; R = randomization; Tx = treatment; RCT = randomized controlled trial; OL = open-label; RW = randomized withdrawal; ABPM = ambulatory blood pressure monitor.

### OUTCOMES

The primary outcome measure in SEQUOIA was designed to capture treatment benefit assessed by OHQ item 1 (dizziness/lightheadedness) ^21^ on ampreloxetine compared to placebo. The primary outcome measure in REDWOOD was designed to capture treatment failure (i.e., worsening) in patients withdrawn to placebo, compared to those maintained on ampreloxetine treatment. Treatment failure was defined as the occurrence of both: 1) a ≥1-point increase (worsening) in symptoms of dizziness/lightheadedness (OHQ item 1) and 2) a ≥1-point increase (worsening) on the Patient Global Impression of Severity (PGI-S) scale. ^23^

Key secondary efficacy outcome measures included individual items and composite scores from the OHQ scale. ^21^ Exploratory endpoints included blood pressure and heart rate measurements obtained from an active stand test (10-minutes supine and 3-minutes standing). Pharmacokinetics and pharmacodynamics were measured at baseline and at the end of the 4-week RCT phase of SEQUOIA.

Clinical laboratory safety tests included vital signs, 12-lead ECG, comprehensive metabolic panel, a complete blood count, and urinalysis. Because a primary safety concern in patients with nOH is exacerbation of supine hypertension, the incidence of supine blood pressure obtained in two ways throughout the trial: 1) from office semi-supine (30-degree angle) blood pressure readings; and 2) from 24-h ambulatory blood pressure monitors worn prior to treatment initiation at baseline, and weeks 1 and week 3 in SEQUIOA. Supine hypertension was graded as absent (systolic ≤139 mmHg, diastolic ≤89 mmHg), mild (systolic: 140-159 mmHg, diastolic 90 – 99 mmHg), moderate (systolic: 160-179 mmHg, diastolic: 100-109 mmHg) or severe (systolic ≥180 mmHg, diastolic ≥110 mmHg). ^24^ The Columbia-Suicide Severity Rating Scale was obtained at key visits in SEQUOIA and REDWOOD. ^25^ The incidence and severity of adverse events (AEs) was captured at each visit. The severity of AEs was subjectively graded by the physician as mild/moderate/severe based on assessment. An independent data safety and monitoring committee reviewed the safety data at predefined intervals and met ad hoc to discuss severe events. All AEs were coded to the preferred terms of the Medical Dictionary for Regulatory Activities (version 24.1). Deaths were adjudicated by an independent committee, who were un-blinded to treatment assignment.

### STATISTICAL ANALYSIS

The SEQUOIA and REDWOOD trials were designed with a minimum enrollment target of 40% for MSA patients. However, the trials were statistically powered based on the full population analysis (PD, MSA and PAF). As a result, the subgroup analysis in MSA patients was not powered to demonstrate statistical significance, and no formal hypothesis testing was planned for this subgroup. All p-values presented here are unadjusted for multiple testing and should be interpreted descriptively.

Disposition is presented for all MSA participants enrolled across both studies according to CONSORT reporting guidelines. ^26^ Baseline characteristics were established using pre-treatment assessments collected at entry into the trial (prior to any study treatment) and are presented for the full analysis set of that study. Given that REDWOOD was specifically designed to evaluate the long-term durability of ampreloxetine in patients with MSA, the efficacy analyses presented in this manuscript focus on the RW period of the REDWOOD study and use its full analysis set. Pharmacodynamic results are reported based on the full analysis set from the SEQUOIA study. The full analysis set for both studies included all MSA patients that had undergone randomization and received at least one dose of their assigned regimen (i.e., intention to treat), and had at least one post-baseline OHSA#1 assessment. Safety analyses for both studies were also reported based on the full analysis set.

The primary endpoint in the REDWOOD trial, treatment failure at Week 6 of the RW period, was analyzed using a logistic regression model. The association between treatment and outcome is expressed as an odds ratio with its corresponding 95% confidence interval. Participants in REDWOOD with missing primary endpoint data were imputed as treatment failures. Changes in OHQ endpoints, blood pressure, and heart rate were analyzed using a mixed model repeated measures analysis. The MMRM model included factors for treatment, visit, treatment-by-visit interaction, and the baseline value as a covariate. Changes in catecholamines are presented descriptively using geometric means, geometric mean standard errors and the geometric mean ratio to prior visit. All statistical analyses were performed using SAS software version 9.4 and SAS/STAT version 15.1 (SAS Institute, Cary, NC, USA).

### STANDARD PROTOCOL APPROVALS, REGISTRATIONS, AND PATIENT CONSENTS

The trials were granted ethical approval by Advarra IRB (registration number 00000971) and by regional ethical standards committees (see eTable 2) and conducted according to the ethical principles of Good Clinical Practice, following the International Council for Harmonization of Technical Requirements for Pharmaceuticals for Human Use Harmonized Tripartite Guideline. Written informed consent was obtained prior to participation.

### DATA AVAILABILITY

Theravance Biopharma (and its affiliates) will not be sharing individual deidentified patient data. The study protocols and statistical analysis plans (SAPs) are available on ClinicalTrials.gov for SEQUOIA (<u>protocol</u>; <u>SAP</u>) and REDWOOD (<u>protocol</u>; <u>SAP</u>). A copy of the most recent protocols and SAPs are available on request.

## RESULTS

### POPULATION

Overall, 73 participants with a confirmed diagnosis of MSA with symptomatic nOH were enrolled into the SEQUOIA (71 participants) and REDWOOD (2 participants) studies (**Figure 1**). The most common reasons for screen failures by the Enrollment Steering Committee were: 1) blood pressure fall on upright tilt not deemed severe enough to account for their symptoms of orthostatic intolerance and 2) a MOCA score at or below the 23-point cut off. The full analysis set included 67 MSA patients for SEQUOIA and 40 MSA patients for REDWOOD.

### BASELINE PRE-TREATMENT CHARACTERISTICS

Eligible MSA patients were on average in their mid-60s (range: 43–80), mostly white, with equal numbers of men and women, and a mean disease duration of 1.6 years (*SD*=1.64 years) (**Tables 1-2**). Twenty-two had the cerebellar phenotype and 44 the parkinsonian form. Clinical characteristics were consistent with mid-to-late stage disease, with manifest diagnostic signs/symptoms of MSA; including a high presence of urogenital dysfunction, dysphagia, dysarthria, recurrent imbalance/falls, gait ataxia, rapid deterioration with early need for walking aid, and poor response to levodopa (**Table 2**). ^1,18^ Eighteen (27%) had used a pressor agent in the past (including midodrine and droxidopa) and 37 (55%) were treatment naïve to any nOH treatments. Fourteen (21%) were treated with a stable dose of fludrocortisone.

**Table 1.** Demographics and Clinical Characteristics at Baseline (Pre-Treatment)

| Measure | Placebo<br>(n=33) | Ampreloxetine<br>(n=34) | All<br>(n=67) |
| --- | --- | --- | --- |
| Male | 20 (60.6%) | 15 (44.1%) | 35 (52.2%) |
| White | 31 (93.9%) | 32 (94.1%) | 63 (94.0%) |
| Not Hispanic or Latino | 31 (93.9%) | 30 (88.2%) | 61 (91.0%) |
| Never Smoker | 23 (69.7%) | 24 (70.6%) | 47 (70.1%) |
| Age (years) | 63.7 (9.61) | 63.3 (8.45) | 63.5 (8.98) |
| NE (pg/ml) | 275.7 (30.05) | 261.0 (22.57) | 268.1 (18.47) |
| BMI (kg/m<sup>2</sup>) | 24.34 (3.93) | 25.47 (4.62) | 24.91 (4.30) |
| OHSA #1 Dizziness Score | 6.55 (1.52) | 6.65 (1.63) | 6.60 (1.57) |
| OHSA Composite Score | 5.27 (1.64) | 5.35 (1.75) | 5.31 (1.69) |
| OHDAS Composite Score | 6.44 (1.98) | 6.44 (2.48) | 6.44 (2.22) |
| OHQ Total Score | 5.84 (1.61) | 5.79 (1.96) | 5.81 (1.8) |
| Supine SBP 10 min (mmHg) | 142.9 (22.83) | 141.24 (20.08) | 142.05 (21.30) |
| Supine DBP 10 min (mmHg) | 87.1 (12.76) | 84.29 (14.84) | 85.63 (13.89) |
| Supine HR 10 min (beats/min) | 69.52 (10.23) | 70.85 (9.6) | 70.22 (9.9) |
| Standing SBP 3 min (mmHg) | 101.97 (27.03) | 96.84 (22.98) | 99.36 (24.98) |
| Standing DBP 3 min (mmHg) | 69.57 (16.53) | 64.65 (17.34) | 67.07 (16.98) |
| Standing HR 3 min (beats/min) | 82.83 (14.50) | 83.10 (13.21) | 82.97 (13.74) |
| Weekly Falls Because of Fainting<sup>a</sup> | 7 (21.2%) | 12 (35.3%) | 19 (28.4%) |
| HADS Anxiety Total Score | 6.48 (4.09) | 6.32 (3.41) | 6.40 (3.73) |
| HADS Depression Total Score | 7.09 (3.96) | 7.76 (3.82) | 7.43 (3.87) |

*Note. NE presented as geometric mean (geometric standard error), other continuous measures presented as mean (SD) and categorical presented as n (%); NE=norepinephrine; BMI=body mass index; OHSA=orthostatic hypotension symptom assessment; OHDAS=orthostatic hypotension daily activity scale; OHQ=orthostatic hypotension questionnaire; SBP=systolic blood pressure; DBP=diastolic blood pressure; HR=heart rate; HADS=Hospital Anxiety and Depression Scale; <sup>a</sup>Non-Motor Symptoms Scale (Score domain 1 question 2, frequency >=often (1/week)).*

**Table 2:** Clinical features of MSA Population Set and Stage of Disease at Entry.

| Domain | Placebo<br>(n=33) | Ampreloxetine<br>(n=34) | All<br>(n=67) |
| --- | --- | --- | --- |
| MSA subtype |  |  |  |
| MSA-C | 8 (24.2%) | 14 (41.2%) | 22 (32.8%) |
| MSA-P | 24 (72.7%) | 20 (58.8%) | 44 (65.7%) |
| Unknown | 1 (3.0%) | 0 | 1 (1.5%) |
| Time since MSA diagnosis (y) <sup>a</sup> | 1.6 (1.72) | 1.5 (1.59) | 1.6 (1.64) |
| Duration of nOH (y) | 2.2 (2.21) | 2.0 (1.56) | 2.1 (1.90) |
| Prior use of any nOH drug <sup>b</sup> | 12 (36.4%) | 18 (52.9%) | 30 (44.8%) |
| Prior use of pressor agent <sup>c</sup> | 8 (24.2%) | 10 (29.4%) | 18 (26.9%) |
| Concomitant use of fludrocortisone | 5 (15.2%) | 9 (26.5%) | 14 (20.9%) |
| Use of dopaminergic medication <sup>d</sup> | 23 (69.7%) | 22 (64.7%) | 45 (67.2%) |
| UMSARS part I | 19.73 (8.92) | 22.12 (6.76) | 20.94 (7.93) |
| UMSARS part II | 22.27 (10.07) | 22.74 (9.16) | 22.51 (9.54) |
| UMSARS part IV |  |  |  |
| *Completely independent* | 3 (9.1%) | 3 (8.8%) | 6 (9.0%) |
| *Not completely independent* | 14 (42.4%) | 14 (41.2%) | 28 (41.8%) |
| *More dependent* | 5 (15.2%) | 7 (20.6%) | 12 (17.9%) |
| *Very dependent* | 10 (30.3%) | 10 (29.4%) | 20 (29.9%) |
| *Totally dependent* | 1 (3.0%) | 0 | 1 (1.5%) |
| Walking aid needed<sup>e</sup> | 21 (63.6%) | 26 (76.5%) | 47 (70.1%) |
| Imbalance/falls<sup>f</sup> | 12 (36.4%) | 15 (44.1%) | 27 (40.3%) |
| Dysarthria<sup>g</sup> | 19 (57.6%) | 22 (64.7%) | 41 (61.2%) |
| Dysphagia<sup>h</sup> | 5 (15.2%) | 10 (29.4%) | 15 (22.4%) |
| Urinary dysfunction<sup>i</sup> | 16 (48.5%) | 18 (58.1%) | 34 (53.1%) |
| Gait impairment<sup>k</sup> | 23 (69.7%) | 27 (79.4%) | 50 (74.6%) |

*Note. Continuous measures are presented as mean (SD) and categorical as n (%); MSA = multiple system atrophy; nOH=neurogenic orthostatic hypotension; UMSARS = Unified Multiple System Atrophy Rating Scale; <sup>a</sup>Time since diagnosis of MSA; <sup>b</sup>Includes vasoactive agents and volume expansion; <sup>c</sup>Includes direct and indirect vasoactive agents midodrine, droxidopa, etilefrine, acarbose, pyridostigmine, atomoxetine, pseudoephedrine; <sup>d</sup>Includes levo-dopa, amatadine, dopamine agonists (pramipexole, ropinirole, rotigotine, piridedil), MAO-B inhibitors (rasagiline, selegiline, safinamide), opicapone, or entacapone; <sup>e</sup>UMSARS question 7 score 2 or above (assistance/walking aid needed occasionally or more); <sup>f</sup>UMSARS question 8 score 2 or above (falling < once a week, more than a week, or cannot walk); <sup>g</sup>UMSARS question 1 speech domain >=2 moderate or worse; <sup>h</sup>UMSARS question 2 swallowing >=2 moderate or worse; <sup>i</sup>UMSARS question 10 >=2 urgency frequency moderate or above requiring drug treatment or catheterization; <sup>k</sup>UMSARS part II question 14 score 2 or above (moderate, severe or cannot walk).*

Prior to receiving ampreloxetine, the mean baseline OHQ item 1 dizziness/lightheadedness score was 6.6 (*SD=*1.57). As shown in **Table 1**, mean systolic blood pressure in the supine position was in the mild supine hypertensive range. Systolic blood pressure fell 43 mmHg within 3 minutes of standing or tilt, with only a minimal increase in heart rate (13 beats/min). All patients met criteria for nOH at entry. ^19^

### 4-WEEK RANDOMIZED CONTROL TRIAL (SEQUOIA)

A total of 71 patients with MSA were randomized and 3 MSA participants dropped out. Over the 4-week double-blind, placebo-controlled, randomized trial with 1:1 allocation, OHQ item 1 scores were not different in the placebo-treated vs. ampreloxetine-treated arms (**eTable 3**).

### 22-WEEK RANDOMIZED WITHDRAWAL TRIAL (REDWOOD)

#### Open label

As shown in the CONSORT diagram, 64 patients with MSA rolled over from SEQUOIA into the REDWOOD trial. Two MSA patients entered de-novo. Of the 66 participants with MSA who entered the REDWOOD open-label phase, 40 (61%) completed the 16-week steady-state treatment, met enrichment criteria for improvement in symptoms, and went on to randomize (**Figure 1**). The main reasons for dropouts during the open label were failure to meet the enrichment continuation criteria (n=8), adverse events (n=7), withdrawal by patient for other reasons including motor deterioration (n=6). At the end of the 16-week open-label, the OHQ item 1 dizziness/lightheadedness score and the nOH Symptom Composite score (OHSA decreased by 3.6 and 2.6 points respectively from the pre-treatment baseline values (**Figure 3B**).

**Figure 3:**
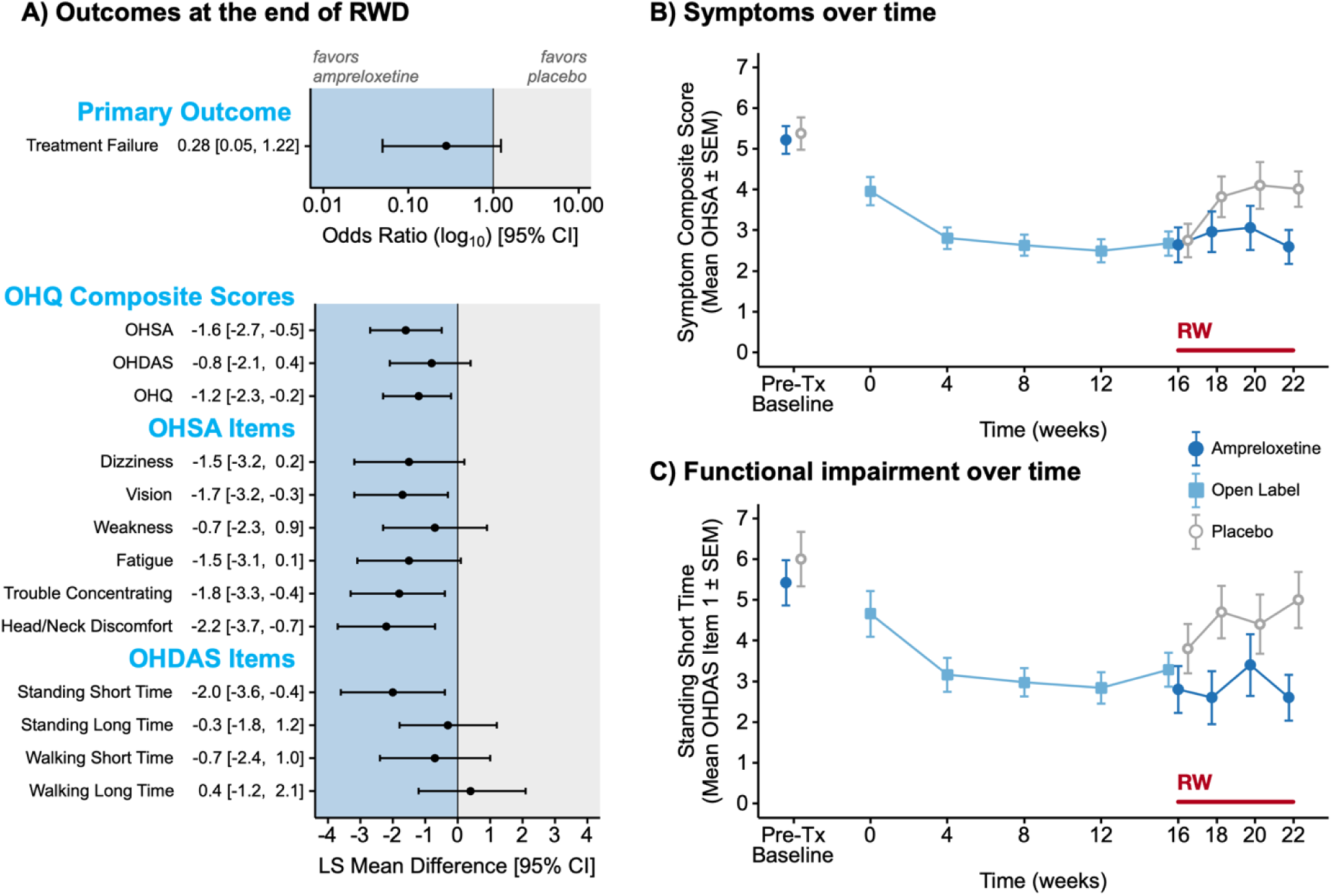
MSA Subgroup Patient-Reported Outcome Measures. A) Forest plot indicates odds of treatment failure (top) and least squares mean difference in OHQ self-reported symptoms scores (bottom) at the end of the 6-week RW. B) Shows longitudinal analysis of mean OHQ symptom assessment composite scores, after censorship for early withdrawal, in the subgroup of randomized MSA patients (n=40). Note, benefit (lower scores) observed in open-label was maintained in the MSA group randomized to ampreloxetine (n=20), but lost (i.e., increasing back to baseline) in the group withdrawn to placebo (n=20). C) Shows OHDAS item 1 scores capturing severity of interference of symptoms of nOH on standing for a short time, which showed a similar pattern with improvement on open-label, that worsening by the end of the randomized withdrawal in the placebo arm. RW = randomized withdrawal; OHQ = orthostatic hypotension questionnaire; OHSA = OH Symptom assessment; OHDAS = OH daily activity scale; LS = least squares; SEM = standard error of the mean; Tx = treatment.

#### Randomized withdrawal

The full analysis set for the pre-specified subgroup analysis of MSA included 20 MSA participants randomized to placebo and 20 randomized to continue 10 mg/day ampreloxetine (MSA, per protocol population). The clinical characteristics at baseline were balanced in the treatment assignment arms in terms of duration of illness, prior use of pressor agents, pre-treatment symptom scores, and UMSARS scores (**Table 2**).

### SYMPTOMS AND DAILY ACTIVITIES

Although the study was not powered to detect differences between the MSA participants, at the end of the RW, the MSA subgroup trended towards a higher rate of treatment failure in those withdrawn to placebo (*OR*=0.28 95% CI [0.05–1.22]) representing a reduction in the odds of treatment failure in ampreloxetine relative to placebo of 72%. Analysis of the OHQ showed individual items on the symptom scale favoring ampreloxetine over placebo at the end of the 6-week RW (**Figure 3A**). This difference was greatest in the least squared (LS) mean difference of the 6-item OHSA symptom burden composite assessment at the end of the RW. The benefit persisted in the MSA patients that continued on ampreloxetine but was lost in the those randomized to withdraw off-treatment to placebo (**Figure 3B**).

At the end of the RW, in addition to symptomatic worsening, the benefit on daily activity scores was lost after 6-weeks in the MSA group assigned to placebo withdrawal. This was most noticeable in OHDAS subscale items that captured short term daily living activities (**Figure 3C**). Walking for a short time (OHDAS item 1) and standing for a short time (OHDAS item 3) both favored continued benefit on ampreloxetine, with the biggest improvements over time observed in patients with lower UMSARS part 4 disability scores. Walking for a long time (OHDAS item 4), was the only individual item not to favor ampreloxetine.

### MECHANISM OF ACTION

As shown in **Table 2**, at entry and prior to randomization in SEQUOIA, pre-treatment venous plasma NE levels in the MSA cohort were 268 pg/ml (range: 74–1014 pg/ml). After 4-weeks of randomization, the catecholamine pharmacodynamic profiles on ampreloxetine showed increased venous plasma NE concentrations compared to both pre-treatment baseline and compared to placebo (**Figure 4A**). Plasma DHPG levels were lowered after 4 weeks on ampreloxetine by 17% (**eTable 3**).

**Figure 4:**
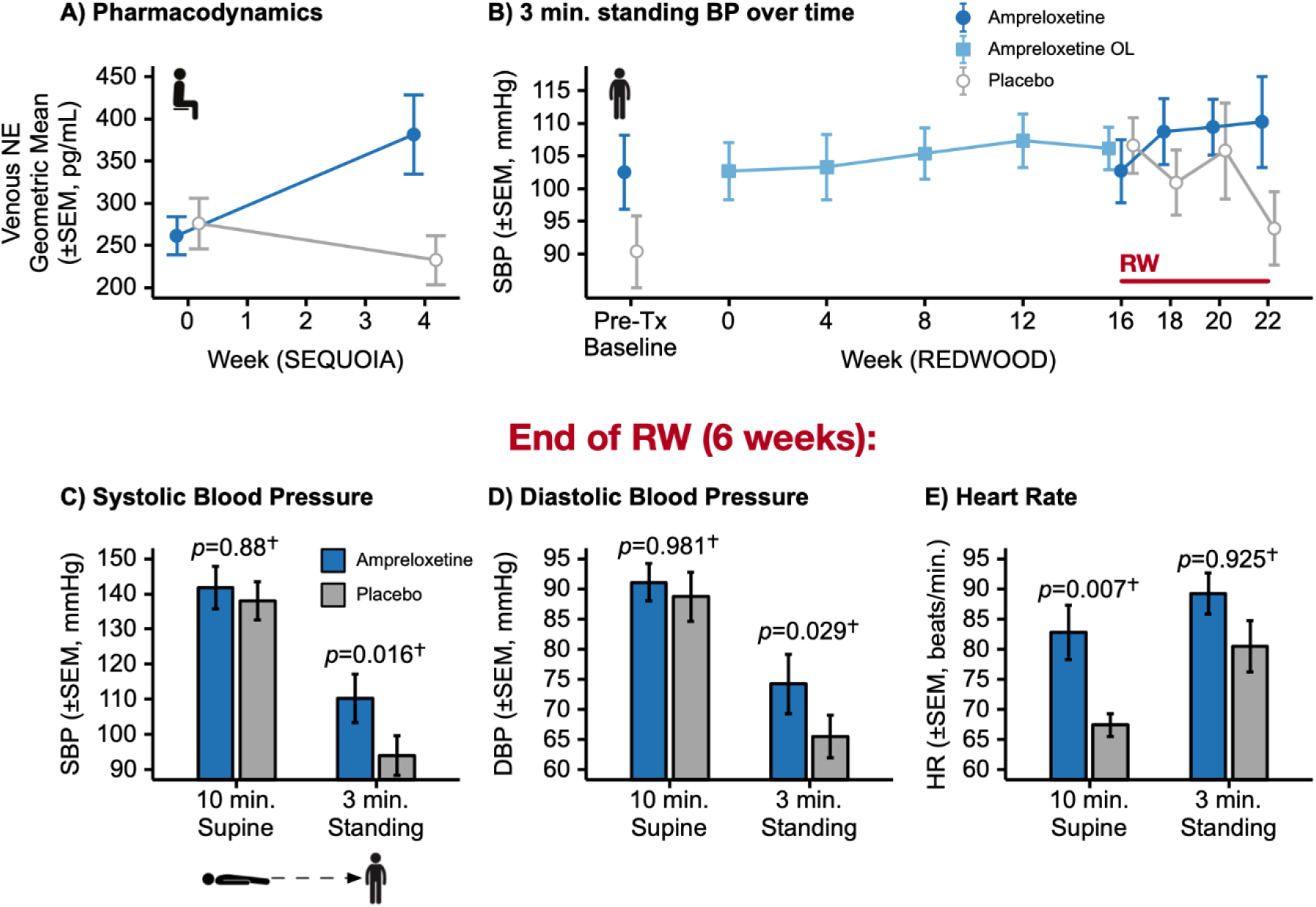
Norepinephrine and Blood Pressure. A) Shows venous plasma NE levels (seated) at baseline and at the end of the 4-week double-blind RCT period of SEQUOIA. B) Shows longitudinal analysis of blood pressure at 3 minutes of standing at entry (Pre-treatment baseline), throughout the 16-week open-label period, and in the randomized withdrawal in the analysis population set of REDWOOD. C) Shows absolute systolic blood pressure after 10 minutes of supine rest and after 3 minutes of standing at week 6 of the RW. D) Shows absolute diastolic blood pressure after 10 minutes of supine rest and after 3 minutes of standing at week 6 of the RW. E) Shows absolute heart rate after 10 minutes of supine rest and after 3 minutes of standing at week 6 of the RW. ^†^P-values vs placebo are based on LS mean difference in change between the start and end of RW. Note, minimal effect of ampreloxetine on supine blood pressure with lower standing blood pressures and heart rate in the placebo group. NE = norepinephrine; RW = randomized withdrawal; SBP = systolic BP; DBP = diastolic blood pressure; HR = heart rate.

**Figure 4B** shows the longitudinal analysis of standing systolic blood pressure throughout the REDWOOD trial. A pressor effect was observed on open-label treatment, with 3 minutes standing blood pressure increasing 6.5 mmHg systolic and 5.3 mmHg diastolic over the 16-week period compared to pre-treatment measures. At the end of the randomized withdrawal, compared to open-label, standing 3-minute blood pressure dropped in the group withdrawn to placebo (systolic: -10.0±4.45, diastolic: -6.0±3.09 mmHg), while it remained stable in group assigned to continue ampreloxetine (systolic: 5.6±4.14; p=0.0157, diastolic: 3.7±2.89; p=0.0293). As a result, at the final RW visit, blood pressure after 3 minutes of standing was higher in the ampreloxetine treated group (**Figure 4C and D**).

### SAFETY AND TOLERABILITY

Ampreloxetine was well-tolerated and there were no safety concerns (**Table 3**). The most common AEs observed in the ampreloxetine group were urinary tract infections and headache. Blood pressures obtained in clinic after 10 minutes supine showed no changes on open-label treatment and in the RW phase in either arm (**Figure 4C**). During SEQUOIA and the RW of REDWOOD there was only 1 AE for supine hypertension which was in the placebo arm during SEQUOIA.

**Table 3:** Safety Analysis.

|  | **SEQUOIA**<br>**(4 week RCT)** |  | **REDWOOD**<br>**(6 weeks RW)** |  |
| --- | --- | --- | --- | --- |
|  | Placebo<br>(n=33) | Amprexetine<br>(n=34) | Placebo<br>(n=20) | Amprexetine<br>(n=20) |
| Subject with any Treatment-Emergent:<br>*Adverse event* | 19 (57.6%) | 19 (54.3%) | 7 (35.0%) | 4 (20.0%) |
| *Moderate or severe AE* | 2 (6.1%) | 9 (25.7%) | 2 (10.0%) | 1 (5.0%) |
| *Severe AE* | 0 | 2 (5.9%) | 1 (5.0%) | 1 (5.0%) |
| *AE related to study drug* | 5 (15.2%) | 5 (14.3%) | 1 (5.0%) | 1 (5.0%) |
| *Serious AE* | 2 (6.1%) | 4 (11.4%) | 1 (5.0%) | 2 (10.0%) |
| *Serious AE related to study drug* | 0 | 0 | 1 (5.0%) | 1 (5.0%) |
| AE leading to permanent discontinuation | 0 | 1 (2.9%) | 0 | 0 |
| Death during study | 0 | 0 | 0 | 0 |
| Subjects with at least 1 AE in >3%<sup>a</sup> | 5 (15.2%) | 8 (22.9%) |  |  |
| *Headache* | 2 (6.1%) | 3 (8.6%) |  |  |
| *Urinary tract infection* | 2 (6.1%) | 3 (8.6%) |  |  |
| *Muscle spasms* | 1 (3.0%) | 2 (5.7%) |  |  |

AE = adverse event. <sup>a</sup>Subjects with at least 1 treatment-emergent AE, by preferred term, which occurred at a frequency of >3% of MSA patients.

## DISCUSSION

The results of this pre-specified subgroup analysis of MSA patients who completed the ampreloxetine phase III trials show a durable symptomatic benefit with improvement in activities of daily living that was lost after ampreloxetine was withdrawn. The pharmacodynamic profile was consistent with the activity of a highly selective peripheral NE transporter inhibitor that increases bioavailability of NE by attenuating NE reuptake at the neurovascular junction, ^16^ with a consequent pressor effect ^27^ and positive ionotrophic/chronotropic effect on heart rate. ^5^

Although the primary outcome was not met, all outcomes measured by the OHQ (with one exception – walking for a long time) including the primary endpoint favored ampreloxetine (See **Figure 3A**). Specifically, the mean between-group differences in the OHQ total composite score, the Orthostatic Hypotension Symptom Assessment (OHSA) composite score, and the Orthostatic Hypotension Daily Activities Scale (OHDAS) composite score all indicated a treatment benefit for ampreloxetine. Importantly, the magnitude of these observed differences met or exceeded the range of minimally important differences previously established through anchor-based analyses in the OHQ validation paper. ^21^ This suggests that the observed benefits of ampreloxetine over placebo across these composite scores is considered clinically meaningful by patients.

The beneficial response observed with ampreloxetine in patients with MSA supports the hypothesis that NE reuptake inhibition is ideally suited to patients with central autonomic lesions, who have relatively intact peripheral autonomic neurons, and normal venous NE levels. Despite the REDWOOD trial being underpowered to show significance in the MSA subgroup analysis, the positive findings indicate a robust durable treatment response in this patient population achieved through selective NET inhibition and support this targeted approach to this specific nOH subpopulation.

A strength of this trial design is that it permits assessment of the durability of clinical efficacy - an important feature in MSA where most patients are likely to require therapy for several years. The results suggest that the reduction in symptom burden (see **Figure 3B**) and the functional impairment burden (see **Figure 3C**) is retained over the 22 weeks of the study. In comparison, the FDA approval for droxidopa was based on pivotal clinical trials of only 2 weeks duration.

The results of this study highlight the challenges of conducting an nOH clinical trial in MSA patients. Several items of the OHQ also may address cardinal non-orthostatic features of MSA related to motor impairment, which may limit their ability to capture the efficacy of an anti-hypotensive drug effect. Despite clarifying instructions on the OHQ, e.g., “symptoms due only to your low blood pressure” and “symptoms that appear on standing,” discriminating OH symptoms from MSA symptoms unrelated to nOH is likely to be demanding for most patients. Unsurprisingly, items such as weakness, standing for a long time and walking for a long time were the least responsive to the intervention. Walking for a long time was the only item not favored by ampreloxetine, and is the OHQ item most likely to be confounded by the progressive extrapyramidal and cerebellar dysfunction in MSA. These challenges should be borne in mind in future nOH trials using the OHQ in central synucleinopathies, especially those in the rapidly progressive MSA.

The inclusion criteria in this study encompassed MSA patients with both possible and probable MSA and the studied patients included patients across clinical disease stages. ^18^ Reports suggest that the median time from disease onset to the requirement of ambulatory aids is approximately 3 years with wheelchair dependency at approximately 5 years. ^28^ Consistent with these reports, 70% of the MSA population enrolled in the ampreloxetine program required walking aids or assistance ambulating. Within this context, the results of the present study suggest that by improving the ability to maintain an upright posture, the functional burden of the disease and quality of life could be improved in MSA patients.

The improvement in symptoms and in most functional measures of orthostatic hypotension was associated with an increase in upright blood pressure which was significant, but modest (see **Figure 4**). A prior positive study reported similar pressor responses and poor concordance between the improvement in symptoms and blood pressure changes. ^29^ The discordance is likely due to the temporal nature of the symptoms which were not recorded daily and assessed with the OHQ by recall over one week versus the timing of the blood pressure measurements, which were acquired at a single office visit. Blood pressure at any given time in a patient with nOH is influenced by multiple factors, including time of day, ambient temperature, hydration/volume status, and post-prandial splanchnic venous pooling ^30^ — all of which confound point-in-time hemodynamic measurements ^31^. Nevertheless, ultimately the primary therapeutic aim when treating nOH is to improve daily function by reducing the burden of symptoms, and not on targeting a specific blood pressure goal. ^27,30^ In this context, raising upright blood pressure to levels just above the lower threshold for cerebral autoregulation may be sufficient to result in a significant improvement in symptoms.

Although 3-min standing blood pressures in the ampreloxetine treated group were significantly greater than in the placebo group at the end of the randomized withdrawal, no worsening of supine hypertension was observed (see **Figure 4, C and D**). This finding is in contrast to studies with the direct α1-adrenoreceptor agonist midodrine and the norepinephrine precursor, droxidopa, in which supine hypertension was observed. The observation may be related to the mechanism of action of ampreloxetine, which increases the availability of physiologically released norepinephrine by inhibiting reuptake at the neurovascular junction, rather than direct stimulation of adrenoreceptors by circulating agonists. In support of this notion is the observation that the drug pyridostigmine, which enhances residual sympathetic outflow also has a low prevalence of supine hypertension. It is most likely that the pressor effect is due to vasoconstriction at the neurovascular junction; however, a contribution of norepinephrine induced chronotropy and inotropy cannot be excluded (**Figure 4E**). This is a topic suitable for further study.

Measurements of venous plasma norepinephrine and metabolites lend support to the study rationale, effective target engagement, and the mechanism of action of ampreloxetine. First, the preservation of peripheral autonomic neurons is supported by the normal venous plasma norepinephrine levels in the patient cohort; second, venous plasma DHPG levels were lower compared to pre-treatment baseline in the ampreloxetine group indicating reduced intraneuronal NE metabolism; and finally, the shift in the DPHG:NE ratio supports target inhibition of the NE transporter and reduced pre-synaptic reuptake at the neurovascular junction. Taken together these neurochemical results provide the basis for the efficacy of ampreloxetine in this patient population.

There are several limitations to our study. First, the trial was not powered to show significance in the MSA subgroup, and all p-values are descriptive. Second, the pre-defined chosen endpoints, as in all nOH studies, were subjective patient reported outcomes. REDWOOD relied on treatment failure as a primary outcome, which equally weighted a 1-point worsening in both the 11-point OHQ item 1 and the 4-point PGI-S scale, ^21,23^ and may have low sensitivity to detect responsiveness in this rapidly progressive neurodegenerative disease. ^31^

In summary, these findings support the ongoing development of ampreloxetine as a targeted treatment for nOH and represent the first example of precision medicine in autonomic neurology and MSA. Based on the results of this pre-specified subgroup analysis, a phase III trial of ampreloxetine focused on patients with MSA (CYPRESS NCT # 05696717) is in progress to confirm these findings.

## Acknowledgments

We would like to thank members of the Enrollment Steering Committee, Investigators and coordinators at recruiting sites as well as patients and their caregivers.

## SUPPLEMENTAL DATA

**eTable 1.** Geographic Location of Randomized Patients With MSA.

|  | Placebo<br>(N=20) | TD-9855 10mg<br>(N=20) | Total<br>(N=40) |
| --- | --- | --- | --- |
| **Europe** | **14 (70.0%)** | **16 (80.0%)** | **30 (75.0%)** |
| Italy | 0 | 6 (30.0%) | 6 (15.0%) |
| Poland | 3 (15.0%) | 1 (5.0%) | 4 (10.0%) |
| Spain | 3 (15.0%) | 1 (5.0%) | 4 (10.0%) |
| Russia | 2 (10.0%) | 1 (5.0%) | 3 (7.5%) |
| United Kingdom | 2 (10.0%) | 1 (5.0%) | 3 (7.5%) |
| France | 0 | 2 (10.0%) | 2 (5.0%) |
| Germany | 1 (5.0%) | 1 (5.0%) | 2 (5.0%) |
| Portugal | 0 | 2 (10.0%) | 2 (5.0%) |
| Austria | 1 (5.0%) | 0 | 1 (2.5%) |
| Bulgaria | 1 (5.0%) | 0 | 1 (2.5%) |
| Denmark | 1 (5.0%) | 0 | 1 (2.5%) |
| Ukraine | 0 | 1 (5.0%) | 1 (2.5%) |
| **North America** | **4 (20.0%)** | **3 (15.0%)** | **7 (17.5%)** |
| United States | 4 (20.0%) | 2 (10.0%) | 6 (15.0%) |
| Canada | 0 | 1 (5.0%) | 1 (2.5%) |
| **Asia/Pacific** | **2 (10.0%)** | **1 (5.0%)** | **3 (7.5%)** |
| Australia | 1 (5.0%) | 1 (5.0%) | 2 (5.0%) |
| Israel | 1 (5.0%) | 0 | 1 (2.5%) |

### eMethods. Full Inclusion/Exclusion/Continuation Criteria for the Trial in the MSA Subgroup

#### Inclusion Criteria at Entry (SEQUOIA)

1. Subject is male or female and at least 30 years old.
2. Subject must meet the diagnostic criteria of nOH, as demonstrated by a sustained reduction in BP of ≥20 mmHg (systolic) or ≥10 mmHg (diastolic) within 3 min of being tilted up to ≥60o from a supine position as determined by a tilt-table test. Subject must score at least a 4 on the Orthostatic Hypotension Symptom Assessment Question #1 at randomization visit.
3. For subjects with MSA only: Subject has a diagnosis of possible or probable MSA of the Parkinsonian subtype (MSA-P) or cerebellar subtype (MSA-C) according to The Gilman Criteria (2008).
4. Subject has plasma NE levels >100 pg/mL after being in seated position for 30 minutes.
5. Subject is willing and able to provide signed and dated written informed consent to participate prior to initiation of any study related procedures.
6. Subject is able to communicate well with the Investigator and clinic staff, understands the expectations of the study and is able to comply with the study procedures, requirements, and restrictions.
7. Subject is female and must be nonpregnant and nonlactating. A woman of childbearing potential must have a documented negative pregnancy test at screening. **NOTE:** A woman is considered to be of childbearing potential unless she is postmenopausal (amenorrheic for at least 2 years) or documented to be surgically sterile (bilateral tubal ligation or total hysterectomy). A female subject may be admitted to the study on the basis of a negative urine pregnancy test. If the urine bHCG (beta human chorionic gonadotropin) test is positive, a serum bHCG test must be performed. The pregnancy test must be confirmed negative for a subject to be eligible for this study.
8. During the study and for 30 days after receiving the last dose of the study drug, females of childbearing potential or males capable of fathering children must agree to use highly effective birth control measures (failure rate <1% when used)

#### Inclusion Criteria into REDWOOD (For SEQUOIA ROLLOVERS)

1. Completion of 4 weeks of double-blind treatment in Study 0169 (V6) and, in the opinion of the Investigator, could benefit from continued treatment with TD-9855. No minimum score of OHSA#1 is required to enter V1 of Study REDWOOD.
2. The subject has a minimum of 80% study medication compliance in Study 0169.
3. The subject must be able to understand the nature of the study and must provide written informed consent prior to the conduct of any study procedures (including an understanding that entry to Study 0170 may result in changes occurring in the subject’s current therapeutic regimen).
4. The subject must be willing to continue on treatment regardless of the possibility of randomization to either TD-9855 or PBO during the randomized withdrawal phase and must continue to meet the inclusion criteria for the preceding study (Study 0169) with the exception that tilt-table test, ESC review and approval of eligibility are not required for entry into Study 0170.

### Exclusion Criteria at Entry (SEQUOIA)

1. Subject has a known systemic illness known to produce autonomic neuropathy, including but not limited to amyloidosis and autoimmune neuropathies. Subject with diabetes mellitus and MSA, will be evaluated on a case by case basis by the medical monitor and considered ineligible unless they meet all of the following criteria:

a. Well controlled type-2 DM in treatment with only oral medications and diet
b. HgbA1C of ≤7.5% performed during screening or up to 12 weeks before screening
c. No clinically evident peripheral neuropathy (e.g., normal sensory examination on peripheral extremities)
d. No known retinopathy (e.g., annual ophthalmic exam is sufficient)
e. No nephropathy (e.g., absence of albuminuria and GFR >60)
2. Subject has a known intolerance to other NRIs or serotonin norepinephrine reuptake inhibitors (SNRIs).
3. Subject currently uses concomitant antihypertensive medication for the treatment of essential hypertension
4. Subject has used strong CYP1A2 inhibitors or inducers within 7 days or 5 half-lives, whichever is longer, prior to randomization or requires concomitant use until the follow-up visit.
5. Subject has changed dose, frequency, or type of prescribed medication for orthostatic hypotension within 7 days prior to randomization visit.

a. Midodrine and droxidopa (if applicable) must be tapered off at least 7 days prior to randomization.
6. Subject has a known or suspected alcohol or substance abuse within the past 12 months (DSM-IV-TR® definition of alcohol or substance abuse).
7. Subject has a clinically unstable coronary artery disease, or major cardiovascular or neurological event in the past 6 months.
8. Subject has used any monoamine oxidase inhibitor (MAO-I) within 14 days prior to randomization.
9. Subject has a history of untreated closed angle glaucoma, or treated closed angle glaucoma that, in the opinion of an ophthalmologist, might result in an increased risk to the subject.
10. Subject has any significant uncontrolled cardiac arrhythmia.
11. Subject has a Montreal Cognitive Assessment (MoCA) ≤23.
12. Subject is unable or unwilling to complete all protocol specified procedures including questionnaires.
13. Subject had a myocardial infarction in the past 6 months or has current unstable angina.
14. Subject has known congestive heart failure (New York Heart Association [NYHA] Class 3 or 4).
15. Subject has any malignant disease other than carcinoma in situ of the cervix or basal cell carcinoma within the past 2 years prior to screening.
16. Subject has a known gastrointestinal (GI) condition, which in the Investigator’s judgment, may affect the absorption of study medication (e.g., ulcerative colitis, gastric bypass).
17. Subject has psychiatric, neurological, or behavioral disorders that may interfere with the ability of the subject to give informed consent or interfere with the conduct of the study.
18. Subject is currently receiving any investigational drug or has received an investigational drug within 30 days of dosing. An investigational drug is defined as nonregulatory agency approved drug (e.g., Food and Drug Administration).
19. Subject has a clinically significant abnormal laboratory finding(s) (e.g., alanine aminotransferase [ALT] or aspartate aminotransferase [AST] >3.0 x upper limit of normal [ULN]; blood bilirubin [total] >1.5 x ULN; estimated glomerular filtration Protocol rate (eGFR) <30 mL/min/1.73m2, or any abnormal laboratory value that could interfere with safety of the subject).
20. Subject has demonstrated a history of lifetime suicidal ideation and/or suicidal behavior, as outlined by the C-SSRS (Baseline/Screening Version) subject should be assessed by the rater for risk of suicide and the subject’s appropriateness for inclusion in the study.
21. Subject has a concurrent disease or condition that, in the opinion of the Investigator, would confound or interfere with study participation or evaluation of safety, tolerability, or pharmacokinetics of the study drug.
22. Subject has known hypersensitivity to TD-9855 (ampreloxetine hydrochloride), or any excipients in the formulation.
23. Subject has:

a. Confirmed severe acute respiratory syndrome coronavirus 2 (SARS-CoV-2) documented with coronavirus disease 2019 [COVID-19] positive test result,
b. Suspected of SARS-CoV-2 infection (clinical features without documented test results two weeks after resolution of symptoms and remains asymptomatic until Day 1)
c. Has been in close contact with a person with known (or suspected) SARS-CoV-2 infection and remains asymptomatic until Day 1.

### Exclusion Criteria into REDWOOD (For SEQUOIA Rollovers)

1. Subject may not be enrolled in another clinical trial (other than exiting Study 0169).
2. Subject has psychiatric, neurological, or behavioral disorders that may interfere with the ability of subjects to give informed consent or interfere with the conduct of the study.
3. Medical, laboratory, or surgical issues deemed by the Investigator to be clinically significant.
4. Uncooperative attitude or reasonable likelihood of non-compliance with the protocol.
5. Subject has a concurrent disease or condition that, in the opinion of the Investigator, would confound or interfere with study participation or evaluation of safety, tolerability, or pharmacokinetics of the study drug.

### Prohibitions and Restrictions

1. Subjects must stop the concomitant use of strong CYP1A2 inhibitors and inducers 7 days or 5 half-lives, whichever is longer, prior to randomization. This restriction applies to concomitant medications, herbal supplements (e.g., St John’s Wort), and ordinary dietary intake.
2. Prescribed medications for OH other than fludrocortisone are prohibited
3. Alpha blockers are prohibited (e.g., Prazosin, Terazosin, Doxazosin, Silodosin, Alfuzosin, Tamsulosin)
4. Norepinephrine reuptake inhibitors (NRIs) and serotonin and norepinephrine reuptake inhibitors (SNRIs) are prohibited − NRIs (e.g., atomoxetine and reboxetine) and SNRIs (e.g., duloxetine, milnacipran, levomilnacipran, venlafaxine, desvenlafaxine)
5. Psychostimulants (e.g., amphetamine, dextroamphetamine, methylphenidate, pemoline) are prohibited
6. Subjects will be requested to refrain from making any significant dietary changes throughout the duration of the study.
7. Subjects should be reminded to maintain an adequate fluid intake during their scheduled visits

### Continuation Criteria in REDWOOD

#### At visit 3, following the initial 4-week OL treatment

1. Subjects must demonstrate a reduction in OHSA#1 of at least 2 points compared to the baseline value, as determined in Study 0169 for subjects entering from Study 0169

#### At visit 6, upon entry into the Double-Blind RW period, following 16-weeks of OL treatment

1. Subject has OHSA#1 score of <7.
2. Subject’s unused OL study medications (10-mg TD-9855 tablets) are returned to site. The subject has a minimum of 80% study medication compliance in OL treatment period.
3. Subjects with excessive deterioration of disease or symptoms during the OL phase and in the opinion of the Investigator, would not benefit from the continual participation in the study.

**eTable 2.** List of clinical trial sites and respective ethics committees/institutional review boards (IRBs)

| **Country** | **Institution Name** | **Ethics Committee** |
| --- | --- | --- |
| Australia | The Royal Melbourne Hospital Neurology Department | Melbourne Health Human Research Ethics Committee |
| Australia | Concord Hospital | Melbourne Health Human Research Ethics Committee |
| Australia | Kingston Center (Monash Health) | Melbourne Health Human Research Ethics Committee |
| Australia | Perron Institute for Neurological and Translational Science | Bellberry Human Research Ethics Committee |
| Austria | Universitätsklinikum Tulln | Ethikkommission der Medizinischen Universität |
| Austria | Medizinische Universität Innsbruck | Ethikkommission der Medizinischen Universität Innsbruck |
| Bulgaria | MHATNP “Sv. Naum” EADClinic of Neurological Diseases for Locomotor Disorders | Ethics Committee for Clinical Trials |
| Canada | Montreal Neurological Institute & Hospital | McGill University Health Center Research Ethics Board (MUHC REB) – Neuroscience and Psychiatry Panel |
| Canada | University Health Network – Toronto Western Hospital | University Health Network Research Ethics Board |
| Canada | University of Calgary | Conjoint Health Research Ethics Board (CHREB) |
| Denmark | Bispebjerg Hospital | De Videnkabsetiske Komiteer for Region Hovedstaden |
| Denmark | Odense Universitetshospital | De Videnkabsetiske Komiteer for Region Hovedstaden |
| Estonia | West Tallinn Central Hospital | Tallinn Medical Research Ethics Committee |
| Estonia | Tartu University Hospital | Tallinn Medical Research Ethics Committee |
| Estonia | East Tallinn Central Hospital | Tallinn Medical Research Ethics Committee |
| Estonia | Astra Team Clinic | Tallinn Medical Research Ethics Committee |
| France | Chu de Nîmes- Hôpital Caremeau | CPP TOURS – Région Centre – Ouest 1<br>Hôpital Bretonneau – CHRU TOURS<br>Groupement d'imagerie médicale |
| Germany | Charite Universitätsmedizin Berlin | Ethik-Kommission der Albert-Ludwigs-Universität Freiburg |
| Germany | Praxis, Dr med. Christian Oehlwein | Ethik-Kommission der Albert-Ludwigs-Universität Freiburg |
| Germany | Charite – Campus Virchow-Klinikum | Ethik-Kommission der Albert-Ludwigs-Universität Freiburg |
| Hungary | Semmelweis Egyetem | Egészségügyi Tudományos Tanács, Klinikai Farmakológiai Etikai Bizottsága |
| Israel | Kaplan Medical Center | Kaplan MC Ethics Committee |
| Israel | Rabin Medical Center | Rabin MC EC Committee |
| Israel | Tel Aviv Sourasky Medical Center | Tel Aviv Sourasky |
| Italy | Fondazione Policlinico Universitario Agostino Gemelli IRCCS / Istituto di Neurologia | Comitato Etico Della Fondazione Policlinico Universitario Agostino Gemelli IRCCS |
| Italy | A.O. Santa Maria | CEAS UMBRIA Presso Regione Umbria |
| Italy | Università di Bologna – Clinical Neurologica Dipartimento di Scienze Neurologiche | CE-A VEC per le Aziende USL dei Bologna e di Imola, |
| Italy | Fondazione IRCCS CA' Granda Ospedale Maggiore Policlinico – U.O. Neurologia | Comitato Etico Milano Area 2 c/o Segreteria Tecnico Scientifica ed Amministrativa |
| Italy | Università degli studi Gabriele D'Annunzio Chieti-Pescara | Comitato Etico delle province di Chieti e Pescara |
| Italy | AOU San Giovanni di Dio e Ruggi d'Aragona | Comitato Etico Campania Sud |
| Italy | Fondazione PTV – Policlinico Tor Vergata | Comitato Etico Indipendente Presso La Fondazione PTV Policlinico Tor Vergata |
| Italy | Azienda Ospedaliero – Universitaria Policlinico Vittorio Emanuele | Comitato Etico Catania 1 (presso l'Azienda Ospedaliera Universitaria Policlinico Vittorio Emanuele di Catania) |
| Italy | Azienda Ospedaliero-Universitaria Pisana- Ospedale S. Chiara, U.O. di Neurologia - Neurofisiopatologia | Comitato Etico Area Vasta Nord Ovest |
| New Zealand | New Zealand Brain Research Institute | Southern Health and Disability Ethics Committee |
| Poland | Specjalistyczne Gabinety Sp. Zo.o | Komisja Bioetyczna przy Śląkiej Izbie Lekarskiej Ul Grazyńskiego |
| Poland | Instytut Zdrowia | Komisja Bioetyczna przy Śląkiej Izbie Lekarskiej |
| Poland | Specjalistyczna Praktyka Lekarska | Komisja Bioetyczna przy Śląkiej Izbie Lekarskiej |
| Poland | ETG Warszawa | Komisja Bioetyczna przy Śląkiej Izbie Lekarskiej |
| Poland | Neuro Care Sp. Zo.o. Sp. Komandytowa | Komisja Bioetyczna przy Śląkiej Izbie Lekarskiej |
| Poland | Karowska Akademia Neurologii Sp. Zo.o. | Komisja Bioetyczna przy Śląkiej Izbie Lekarskiej |
| Poland | PRATIA MCM Kraków | Komisja Bioetyczna przy Śląskiej Izbie Lekarskiej |
| Portugal | Campus Neurológico Sénior | Comissão de Ética para a Investigação Clínica |
| Portugal | Hospital Senhora da Oliveira – Guimarães, EPE | Comissão de Ética para a Investigação Clínica |
| Russia | Saint Petersburg State Budgetary Institution of Healthcare “City Hospital #40 of Kurkotnyi region” | Ethical Council at the Ministry of Health of Russian Federation, Ethics Council at Saint Petersburg State Budgetary Institution Healthcare “City Hospital #40 of Kurortnyi region” |
| Russia | Limited Liability Company “City Neurological Center Sibneiromed” | Ethical Council at the Ministry of Health of Russian Federation Local Ethics Committee at LLC “City Neurological Center Sibneiromed |
| Russia | Federal State Budgetary Educational Institution of Additional Professional Education “Russian Medical Academy of Continuous Postgraduate Education” | Ethical Council at the Ministry of Health of Russian Federation Ethics Committee at FSBEI of APE” Russian Medical Academy of Continuous Postgraduate Education” of the Ministry of Healthcare of the Russian Federation |
| Russia | Federal State Budgetary Institution “National Medical Research Centre of Psychiatry and Neurology named after VM. Bekhterev” | Ethical Council at the Ministry of Health of Russian Federation Independent Ethics Committee at Federal State Budgetary Institution “National Medical Research Centre of Psychiatry and Neurology named after V.M. Bekhterev” of the Ministry of Healthcare of the Russian Federation |
| Russia | Federal State Budgetary Institution “Federal Siberian Scientific and Clinical Center of Federal Medico-Biological Agency” | Ethical Council at the Ministry of Health of Russian Federation<br>Ethics Committee at Federal State Budgetary Institution “Federal Siberian Scientific and Clinical Center of Federal Medico-Biological Agency” |
| Russia | Federal State budgetary Institution of Science Institute of Human Brain named after N.P. Bekhtereva of Russian Academy of Sciences | Ethical Council at the Ministry of Health of Russian Federation<br>Ethics Committee at FSBI of Science Institute of Human Brain named after N.P. Bekhtereva of Russian Academy of Sciences |
| Russia | State Budgetary Institution of Healthcare of Novosibirsk region<br>“City Clinical Hospital #34” | Ethical Council at the Ministry of Health of Russian Federation |
| Spain | Complejo Hospitalario de Navarra | CEIm Provincial de Sevilla |
| Spain | Hospital Universitario de la Princesa | CEIm Provincial de Sevilla |
| Spain | Hospital Universitario Mutua de Terrasa | CEIm Provincial de Sevilla |
| Spain | Hospital del Mar | CEIm Provincial de Sevilla |
| Spain | Hospital de Cruces | CEIm Provincial de Sevilla |
| UK | Kings College Hospital NHS Foundation Trust | NHSBT Newcastle Blood Donor Centre |
| UK | The National Hospital for Neurology and Neurosurgery,<br>Autonomic Unit | Wales Research Ethics |
| UK | Cognition Health Ltd. | North East – Newcastle & North Tyneside 1 Research Ethics Committee |
| UK | Clinical Research Centre<br>William Harvey Heart Centre,<br>Barts and the London | North East – Newcastle & North Tyneside 1 Research Ethics Committee |
| UK | Royal Devon And Exeter NHS Foundation Trust | North East – Newcastle & North Tyneside 1 Research Ethics Committee |
| UK | Salford Royal NHS Foundation Trust | R&D - Salford Care Organisation |
| UK | Re:Cognition Health Ltd | NRES Committee North East - Newcastle & North Tyneside 1 |
| Ukraine | Communal Noncommercial Enterprise City Polyclinic #9 of Kharkiv City Council | The Ethics Commision at Communal Noncommercial Enterprise "City Polyclinic #9" of Kharkiv City Council |
| Ukraine | Communal Institution Acad. O.I. Yushchenko Vinnytsia Regional Psychoneurological Hospital | The Ethics Commision at Communal Institution Acad O.I. Yushchenko Vinnytsia Regional Psychoneurological Hospital |
| Ukraine | Communal Noncommercial Enterprise of Lviv Regional Council "Lviv Regional Clinical Hospital" | Communal Noncommercial Enterprise of Lviv Regional Council "Lviv Regional Clinical Hospital Department of Neurology |
| USA | Rocky Vista University, Colorado | Quorum Review IRB |
| USA | Selkirk Neurology, PLLC | Quorum Review IRB 1 |
| USA | NorthShore University Health System | Quorum Review IRB |
| USA | The University of Texas Southwestern Medical Center | Quorum Review IRB |
| USA | University of Cincinnati Academic Health Center | University of Cincinnati IRB |
| USA | SFM Clinical Research, LLC | Advarra IRB |
| USA | Oregon Health & Science University (OHSU) | Quorum Review IRB |
| USA | Parkinson's Disease and Movement Disorders Center of Boca Raton | Quorum Review IRB |
| USA | Rush University Medical Center | Rush University Medical Center IRB, Office of Research Affairs |
| USA | UC San Diego Health | UCSD Human Research Protections Program |
| USA | Mayo Clinic | Mayo Clinic IRB |
| USA | NYU Langone Health | NYU Grossman School of Medicine IRB |
| USA | Neurostudies Inc | Quorum Review IRB |
| USA | University of Kansas Medical Center | University of Kansas Medical Center IRB |
| USA | Vanderbilt University Medical Center | Vanderbilt Institutional Review Board |
| USA | Banner Sun Health Research Institute | Western IRB |
| USA | Stanford University Medical Center | Stanford IRB Research and Compliance Office |
| USA | Georgetown University Hospital | Georgetown University IRB |
| USA | Henry Ford Medical Center - West Bloomfield | Henry Ford Health Systems IRB |
| USA | Wake Forest University Baptist Health Sciences | Wake Forest University Health Sciences IRB |
| USA | Ohio State University - Wexner Medical Center | Ohio State University Comprehensive Cancer Center Clinical Scientific Review Committee |
| USA | Colorado Springs Neurological Associates, PC | Quorum Review IRB |

**eTable 3.**
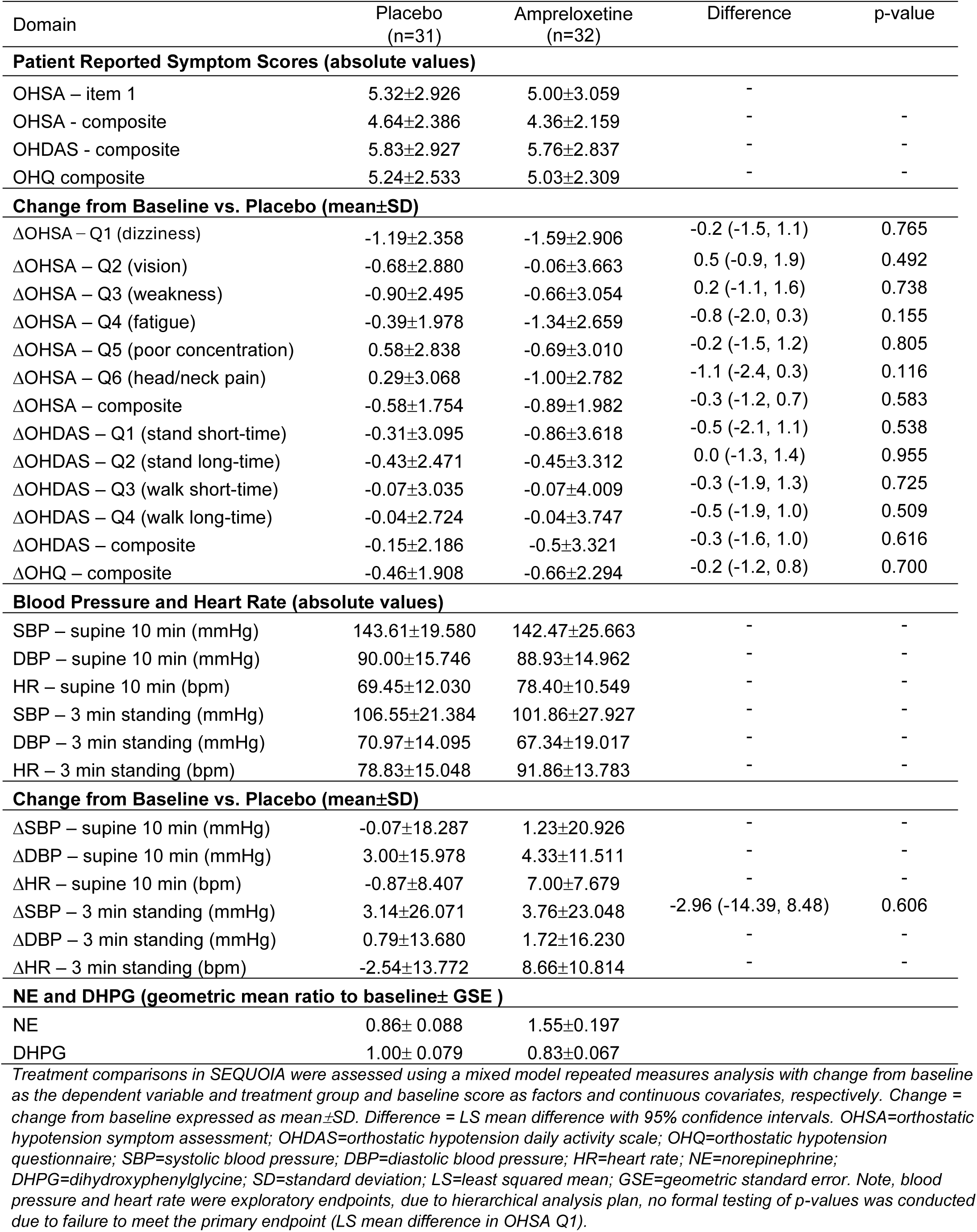
Topline Results, MSA Subgroup, 4-Week RCT (SEQUOIA)

| Domain | Placebo<br>(n=31) | Amprexetine<br>(n=32) | Difference | p-value |
| --- | --- | --- | --- | --- |
| **Patient Reported Symptom Scores (absolute values)** |  |  |  |  |
| OHSA – item 1 | 5.32±2.926 | 5.00±3.059 | - | - |
| OHSA - composite | 4.64±2.386 | 4.36±2.159 | - | - |
| OHDAS - composite | 5.83±2.927 | 5.76±2.837 | - | - |
| OHQ composite | 5.24±2.533 | 5.03±2.309 | - | - |
| **Change from Baseline vs. Placebo (mean±SD)** |  |  |  |  |
| ΔOHSA – Q1 (dizziness) | -1.19±2.358 | -1.59±2.906 | -0.2 (-1.5, 1.1) | 0.765 |
| ΔOHSA – Q2 (vision) | -0.68±2.880 | -0.06±3.663 | 0.5 (-0.9, 1.9) | 0.492 |
| ΔOHSA – Q3 (weakness) | -0.90±2.495 | -0.66±3.054 | 0.2 (-1.1, 1.6) | 0.738 |
| ΔOHSA – Q4 (fatigue) | -0.39±1.978 | -1.34±2.659 | -0.8 (-2.0, 0.3) | 0.155 |
| ΔOHSA – Q5 (poor concentration) | 0.58±2.838 | -0.69±3.010 | -0.2 (-1.5, 1.2) | 0.805 |
| ΔOHSA – Q6 (head/neck pain) | 0.29±3.068 | -1.00±2.782 | -1.1 (-2.4, 0.3) | 0.116 |
| ΔOHSA – composite | -0.58±1.754 | -0.89±1.982 | -0.3 (-1.2, 0.7) | 0.583 |
| ΔOHDAS – Q1 (stand short-time) | -0.31±3.095 | -0.86±3.618 | -0.5 (-2.1, 1.1) | 0.538 |
| ΔOHDAS – Q2 (stand long-time) | -0.43±2.471 | -0.45±3.312 | 0.0 (-1.3, 1.4) | 0.955 |
| ΔOHDAS – Q3 (walk short-time) | -0.07±3.035 | -0.07±4.009 | -0.3 (-1.9, 1.3) | 0.725 |
| ΔOHDAS – Q4 (walk long-time) | -0.04±2.724 | -0.04±3.747 | -0.5 (-1.9, 1.0) | 0.509 |
| ΔOHDAS – composite | -0.15±2.186 | -0.5±3.321 | -0.3 (-1.6, 1.0) | 0.616 |
| ΔOHQ – composite | -0.46±1.908 | -0.66±2.294 | -0.2 (-1.2, 0.8) | 0.700 |
| **Blood Pressure and Heart Rate (absolute values)** |  |  |  |  |
| SBP – supine 10 min (mmHg) | 143.61±19.580 | 142.47±25.663 | - | - |
| DBP – supine 10 min (mmHg) | 90.00±15.746 | 88.93±14.962 | - | - |
| HR – supine 10 min (bpm) | 69.45±12.030 | 78.40±10.549 | - | - |
| SBP – 3 min standing (mmHg) | 106.55±21.384 | 101.86±27.927 | - | - |
| DBP – 3 min standing (mmHg) | 70.97±14.095 | 67.34±19.017 | - | - |
| HR – 3 min standing (bpm) | 78.83±15.048 | 91.86±13.783 | - | - |
| **Change from Baseline vs. Placebo (mean±SD)** |  |  |  |  |
| ΔSBP – supine 10 min (mmHg) | -0.07±18.287 | 1.23±20.926 | - | - |
| ΔDBP – supine 10 min (mmHg) | 3.00±15.978 | 4.33±11.511 | - | - |
| ΔHR – supine 10 min (bpm) | -0.87±8.407 | 7.00±7.679 | - | - |
| ΔSBP – 3 min standing (mmHg) | 3.14±26.071 | 3.76±23.048 | -2.96 (-14.39, 8.48) | 0.606 |
| ΔDBP – 3 min standing (mmHg) | 0.79±13.680 | 1.72±16.230 | - | - |
| ΔHR – 3 min standing (bpm) | -2.54±13.772 | 8.66±10.814 | - | - |
| **NE and DHPG (geometric mean ratio to baseline± GSE )** |  |  |  |  |
| NE | 0.86± 0.088 | 1.55±0.197 |  |  |
| DHPG | 1.00± 0.079 | 0.83±0.067 |  |  |

*Treatment comparisons in SEQUOIA were assessed using a mixed model repeated measures analysis with change from baseline as the dependent variable and treatment group and baseline score as factors and continuous covariates, respectively. Change = change from baseline expressed as mean±SD. Difference = LS mean difference with 95% confidence intervals. OHSA=orthostatic hypotension symptom assessment; OHDAS=orthostatic hypotension daily activity scale; OHQ=orthostatic hypotension questionnaire; SBP=systolic blood pressure; DBP=diastolic blood pressure; HR=heart rate; NE=norepinephrine; DHPG=dihydroxyphenylglycine; SD=standard deviation; LS=least squared mean; GSE=geometric standard error. Note, blood pressure and heart rate were exploratory endpoints, due to hierarchical analysis plan, no formal testing of p-values was conducted due to failure to meet the primary endpoint (LS mean difference in OHSA Q1).*

